# DiaBench: A Benchmarking Framework to Evaluate the Performance of LLMs in T2D Patient Education

**DOI:** 10.64898/2026.10.26.26366562

**Authors:** Yang Li, Syna Syna, Chuyi Zhang, Bhavana Kunisetty, Elizabeth Healey, Agatha F. Scheideman, Mandy M. Shao, Han Meng, Yu Kuei Lin, Jacob Kohlenberg, Yangfu Zhang, David C. Klonoff, Anna Simos, Marina Basina, Michael P. Snyder, Haipeng Chen, Tao Wang

**Author notes:** **Co-corresponding:** Michael Snyder; Haipeng Chen; Tao Wang. Equal contribution (Yang Li, Syna Syna, Chuyi Zhang).

## Abstract

**Background:** Effective type 2 diabetes (T2D) education improves patient outcomes, but clinician time constraints limit what can be covered during encounters. Large language models (LLMs) offer a scalable way to answer patient questions, yet their accuracy and reliability in T2D education across health literacy levels remain poorly characterized.

**Methods:** We developed DiaBench, a standardized benchmarking framework built from 96 question-and-answer pairs collected by clinicians from representative real-world patient encounters. GPT-5 transformed each pair into multiple-choice questions (MCQs) under three configurations (1-3) simulating different question formats and levels of user background. A five-member panel of certified diabetes care and education specialists (CDCESs), medical doctors (MDs), and endocrinologists validated the transformations, rating question rephrasing, answer rephrasing, and answer correctness. We then evaluated seven LLMs (GPT-5^1^, Gemini-2.5-Flash^2^, Cohere Command^3^, Llama-4^4^, DeepSeek-R1^5^, Perplexity Sonar^6^, Mistral-Large-2.0^7^) for accuracy and self-consistency across three runs.

**Results:** (i) Expert validation showed a consensus over MCQ transformation. For configuration 1, 98.1% (SD 0.01) of rephrased answers were appropriate, 98.1% (SD 0.01) of MCQ answers were correct, and 95.0% (SD 0.05) of rephrased questions were appropriate; corresponding values for configuration 2 were 96.3% (SD 0.05), 97.5% (SD 0.02), and 89.8% (SD 0.14). (ii) across benchmarking for configurations 1 and 2, most models achieved high accuracy (≥95%), but Mistral-Large-2.0, Llama 4, and Perplexity Sonar showed substantially lower self-consistency (60.4%-91.7%) than other models (≥95%). All models degraded in both accuracy and self-consistency on configuration 3.

**Conclusions:** DiaBench quantitatively differentiates LLM capability in T2D patient education, with Gemini, GPT-5, and DeepSeek-R1 performing strongly and other models suboptimally. DiaBench provides a transferable platform for evaluating existing and newly developed LLMs, addressing the need for standardized benchmarking before clinical deployment of LLM-based systems in patient education.

## Introduction

Diabetes mellitus remains a rapidly growing health challenge. Effective chronic disease management is complex and requires patient engagement and education^8^. Patients often have questions about their condition, dietary choices, and treatment options. Providing timely answers to patients’ questions about diabetes management has been shown to enhance patient engagement and improve health outcomes ^8^. However, a critical gap exists between these informational needs and the limited time clinicians have to address them adequately^9–12^. Specifically, there is a need for a scalable tool that can deliver reliable information to support T2D patients with varying levels of health literacy^13,14^.

The rapid advancement of large language models (LLMs), driven by innovations in scale and architecture, has demonstrated immense potential across various healthcare applications^15–22^. These tools can extract crucial information from unstructured clinical notes^23–25^ and answer patients’ questions^26–29^. In the field of diabetes care, LLMs are being explored to develop personalized treatment strategies and intelligent question-and-answer (Q&A) systems^14,25,30^. However, a significant barrier to their clinical adoption is the models’ known tendency for hallucination and inconsistent generation^31–33^. This limitation is especially problematic in T2D education, where providing inaccurate information can lead to incorrect or delayed interventions, patient mistrust of providers’ recommendations, and adverse patient outcomes.

Therefore, rigorous evaluation of LLMs is needed to ensure their safe integration into T2D education, as these generalist models are not specialized to address diabetes-related questions. Prominent general benchmarks, such as MedQA^34^, MedMCQA^35^, and PubMedQA^36^, provide structured, multiple-choice frameworks for evaluating LLM performance in understanding medical knowledge. While valuable, these benchmarks are not well-suited to the specific needs of diabetes education because they lack domain-specific questions and are not designed to accommodate patients across a spectrum of health literacy levels^37–39^.

Recognizing this gap, researchers have evaluated LLMs using free-form questions tailored to education for individuals living with diabetes^14,40–43^. However, these efforts rely on manual assessment by clinical experts who rate responses using subjective criteria. Such a manual assessment process is inherently labor-intensive and time-consuming, and thus cannot serve as a reusable benchmark that can be easily rerun to evaluate new LLM models or existing models with different configurations. More recent work has explored the feasibility of using LLMs as judges to replace manual assessment in general domains, but has found this approach to be less reliable than human expert consensus^44–46^. Ultimately, current methods lack a standardized infrastructure required to systematically and objectively evaluate LLMs in the context of T2D education.

To address these gaps, we developed DiaBench, a scalable, standardized MCQ-based benchmark designed to assess LLM performance in T2D education, providing a platform with 3 configurations representing different literacy levels. This allows for the evaluation of both existing and newly developed LLMs. In this study, we utilize DiaBench to conduct a comprehensive, zero-shot evaluation of seven state-of-the-art LLMs, assessing their clinical accuracy and self-consistency to determine their readiness for real-world deployment.

## Methods

In Figure 1, we present the overview of the MCQ curation and evaluation pipelines. First, we discuss the MCQ curation pipeline, which begins with CDCESs, MDs, and endocrinologists collecting an initial Q&A dataset. Second, we discuss the GPT-5-based MCQ transformation and expert assessment. Third, we outline the design and implementation principles underlying the DiaBench LLM evaluation infrastructure.

**Figure 1.**
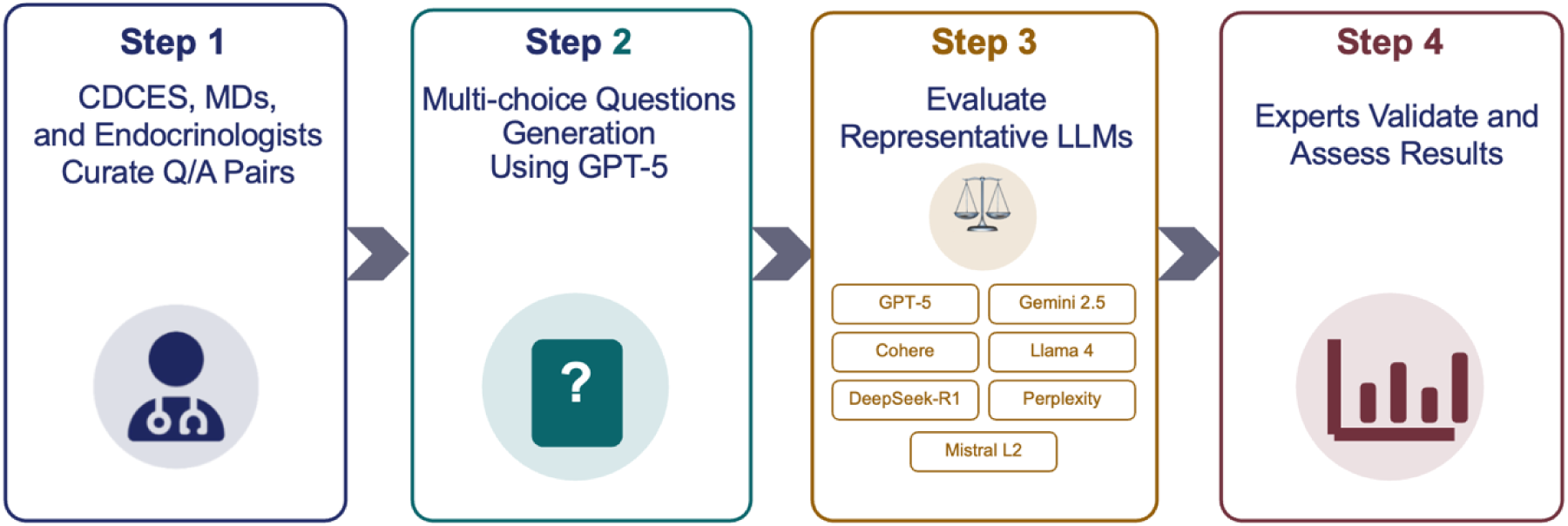
Overview of the DiaBench pipeline from seed question to evaluation process.

### Collecting Initial Question-and-Answer Pairs

DiaBench began by collecting 96 representative frequently asked questions and their free-form answers from CDCESs, MDs, and endocrinologists. These questions represent daily clinician-patient interactions, and their corresponding answers were derived from the most current ADA guidelines. Overall, the initial Q&A pairs collected ensure that the benchmark reflects real-world patient needs. We inspected the selected Q/A pairs and classified them into five categories. If multiple categories may apply, only the most relevant one was chosen. The categories include:

1. Lifestyle modifications, including diet, sleep, and exercise (e.g., “Can I eat bread if I have type 2 diabetes?”)
2. General understanding of the pathophysiology of Type 2 Diabetes (e.g., “Can diabetes be reversed?”)
3. Complications and monitoring (e.g., “Can T2D lead to kidney problems?”)
4. Medical management and treatment for long-term diabetic care (e.g., “What medications are commonly prescribed for T2D?”)
5. Mental and emotional health awareness in people living with diabetes (e.g., “How does T2D affect mental health?”)

### Automated Transformation of MCQs

DiaBench adopted the MCQ format used by existing general LLM benchmarks ^35,36^. By transforming Q&As to the MCQ format, DiaBench can be easily adopted by any existing or under-development LLM without requiring additional human expert evaluations since evaluation is a simple pattern-matching to the correct option(s). On the other hand, the number of the Q&As can be easily scaled up by physicians with no technical hurdles.

The transformation is achieved by prompting GPT-5, one of the top-performing models at the time of research, with a detailed prompting template for 3 configurations. We describe the details in the following sections.

### Different Configuration Considerations

Although the seed questions represent common patient-provider interactions, the original free-text answers did not account for patients’ diverse health and language literacy levels. Ideally, LLMs should be able to provide responses tailored to patients across different literacy levels. To evaluate the impact of linguistic and health literacy levels on LLMs, we used GPT-5 to systematically transform each initial Q&A into MCQs organized into three configurations that represent different user groups. We achieved this by modulating the simulated patient background, the total number of options, and the number of correct choices in the following configurations:

● **Configuration 1:** 9th-grade literacy level; 1 correct answer out of 4 options.
● **Configuration 2:** Undergraduate literacy level; 1 correct answer out of 7 options.
● **Configuration 3:** Graduate literacy level; 2 correct answers out of 10 options.

To quantitatively validate these targeted literacy levels, we applied the Flesch-Kincaid Grade Level^47^, an empirically validated readability metric derived from average sentence length and the average number of syllables per word. A detailed mapping between the raw score and readability level is provided in Appendix A. To further compare readability metrics across the different configurations, we conducted Student’s *t*-tests and Wilcoxon rank-sum tests.

### Prompting Template for MCQ Transformation

To fully leverage GPT-5’s instruction-following and generative capabilities for MCQ transformation, we designed a unified prompt template across all 3 configurations. As indicated in Table 1, the prompt consists of task instructions, configuration requirements, input Q&A format, and output MCQ format. More detailed templates are provided in Appendix B. The generation pipeline was implemented in Python. All function calls to GPT-5 use publicly available OpenAI APIs with default settings, can be easily adapted, and are fully automated. The automated MCQ transformation takes only several minutes to complete.

**Table 1.** Major components of the MCQs transformation prompt.

|  |  |
| --- | --- |
| Task Instruction | Transform a clinician-curated Q&A -> MCQs |
| Configuration Requirements | Reading level, number of options, number of correct answers, ... |
| Input Q&A | The original Q&A pair. |
| Output MCQs Format | The MCQs in the desired format for parsing. |

### Expert Verification Process

As LLMs are known to suffer from hallucinations, a verification procedure by domain experts is needed to ensure the quality of the generated MCQ Q&As. A panel of clinical experts performed a validity check based on the following criteria: (1) Is the generated question a proper rephrasing of the original question? (2) Do the generated answers provide a proper rephrasing of the original answer? (3) Are the generated correct answer(s) the appropriate answer(s) to this question? These criteria are intended to capture semantic distortion but can also catch mistakes such as factual errors, logical fallacies, question-and-answer mismatches, and more.

### Choice of LLMs

We evaluated seven LLMs across open- and closed-sourced top-performing models, selecting those available to the general public (if an API or front-end is available): (1) GPT-5^1^, (2) Gemini 2.5 flash^2^, (3) Cohere command^3^, (4) LLama 4^4^, (5) Deepseek-R1^5^, (6) Perplexity-sonar^6^, and (7) Mistral large 2.0^7^. All models are prompted under a zero-shot setting. For each question, we ran each LLM 3 times to account for the stochasticity in LLM generations, unless otherwise specified.

### Evaluation Pipeline and Performance Metrics

The evaluation pipeline is implemented in Python and uses publicly available APIs, which can be easily adapted to evaluate any desired LLMs with available APIs. We evaluated all models in a strict zero-shot (i.e., no examples given) setting via their respective APIs. To prevent context changes, we passed each question to the model as an isolated, independent query. Once we receive the generated response, we use a Python-based parser to analyze the final results and compare them with the answer labels from previous steps. The full evaluation pipeline is fully automated and can be easily adapted to existing and future LLMs. While our paper focuses on T2D education, this pipeline can be applied to other domains, which is much more scalable than the previous human expert-based evaluation pipeline^14,40^. For each LLM, the entire evaluation pipeline took only several minutes to run.

Regarding the evaluation metrics, we used accuracy and self-consistency throughout the pipeline. Because LLM generations use stochastic sampling, we ran each LLM three times with identical prompts and calculated the accuracy by majority voting across these three independent runs. We defined self-consistency as the model’s ability to generate identical final answers across all three independent runs.

## Results

### Initial Q&A Pairs Distribution

In Figure 2, we present the distribution of the five categories of frequently asked questions collected from real-world encounters, along with their free-form answers from CDCESs, MDs, and endocrinologists. The questions are roughly evenly distributed across categories, with fewer questions from the mental & emotional health category.

**Figure 2.**
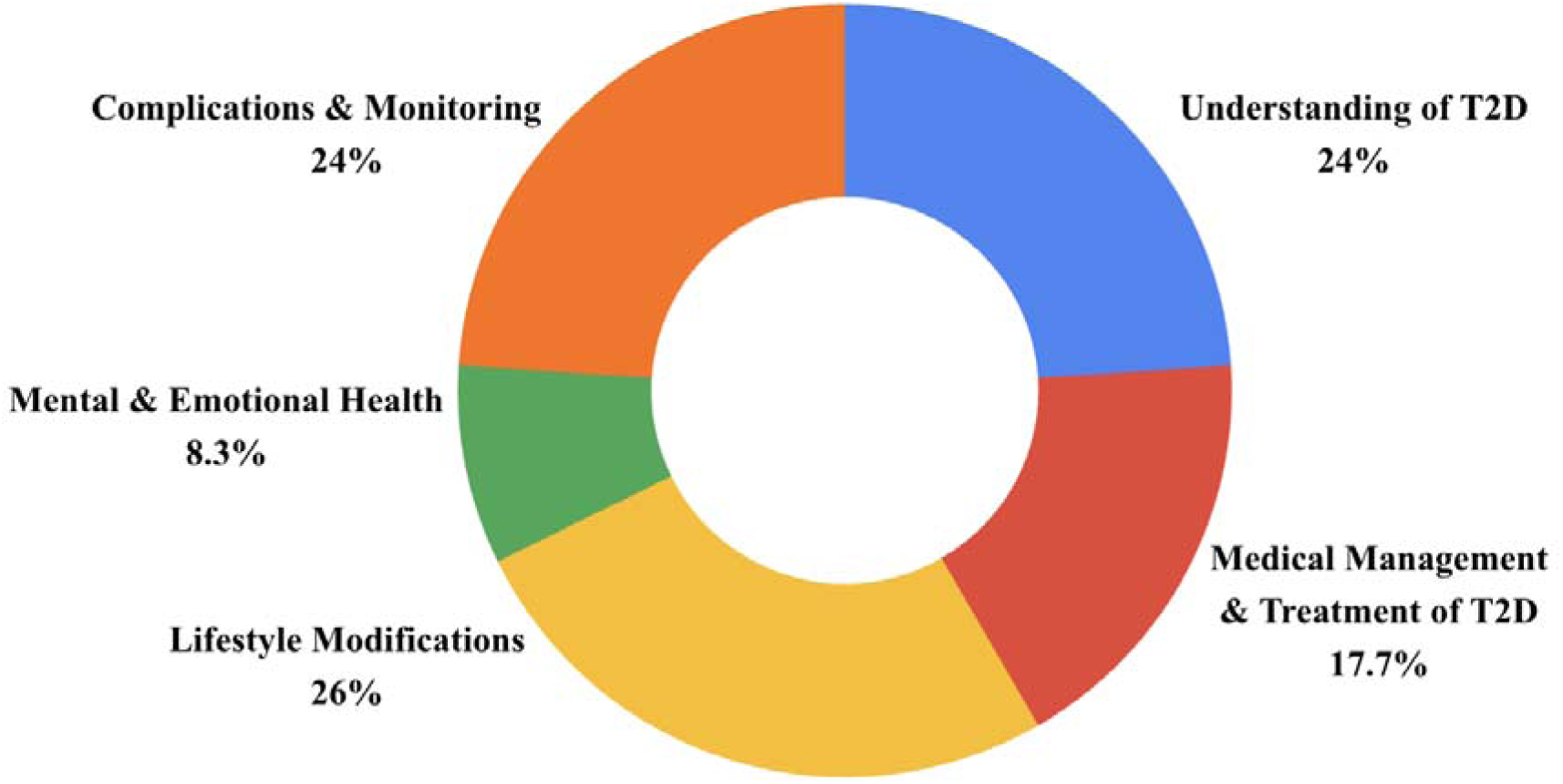
The distribution of the 96 original Q&As collected.

### Verification by Clinical Experts

A panel of CDCESs, MDs, and endocrinologists conducted a rigorous clinical review of the questions generated from configurations 1 and 2. The experts reached a high degree of consensus on the appropriate rephrasing of MCQ answer options and the correctness of generated answers. For configuration 1, verification results showed an appropriate answer rephrasing rate of 98.1% (SD: 0.01), a generated answer correctness rate of 98.1% (SD: 0.01), and an appropriate question rephrasing rate of 95.0% (SD: 0.05). Configuration 2 yielded an appropriate answer rephrasing rate of 96.3% (SD: 0.05), a generated answer correctness rate of 97.5% (SD: 0.02), and an appropriate question rephrasing rate of 89.8% (SD: 0.14). CDCESs, MDs, and endocrinologists reached a consensus to skip configuration 3, as it is usually much more complex and less likely to appear in real-world clinical encounters. Nevertheless, a small subset of the transformed questions was flagged as inappropriate. We will discuss these flagged instances in the next section.

### Flesch-Kincaid Grade Level Verification

The Flesch-Kincaid Grade distribution across the 3 configurations is presented in Figure 3, with higher scores indicating greater required reading proficiency. The mean Flesch-Kincaid scores were 8.66 for Configuration 1, 12.64 for Configuration 2, and 14.69 for Configuration 3. One-sided student’s t-test and Wilcoxon rank-sum results support the hypothesis that configurations 2 and 3 yielded higher Flesch-Kincaid grade scores than configuration 1, with significant p-values (p < 0.001 and p<0.001, respectively). At the same time, such a relationship cannot be established between configurations 3 and 2 for the t-test with p = 0.12 and the Wilcoxon rank-sum test with p = 0.06, respectively. Figure 4 shows the number of questions that fall into the readability score band from each configuration.

**Figure 3.**
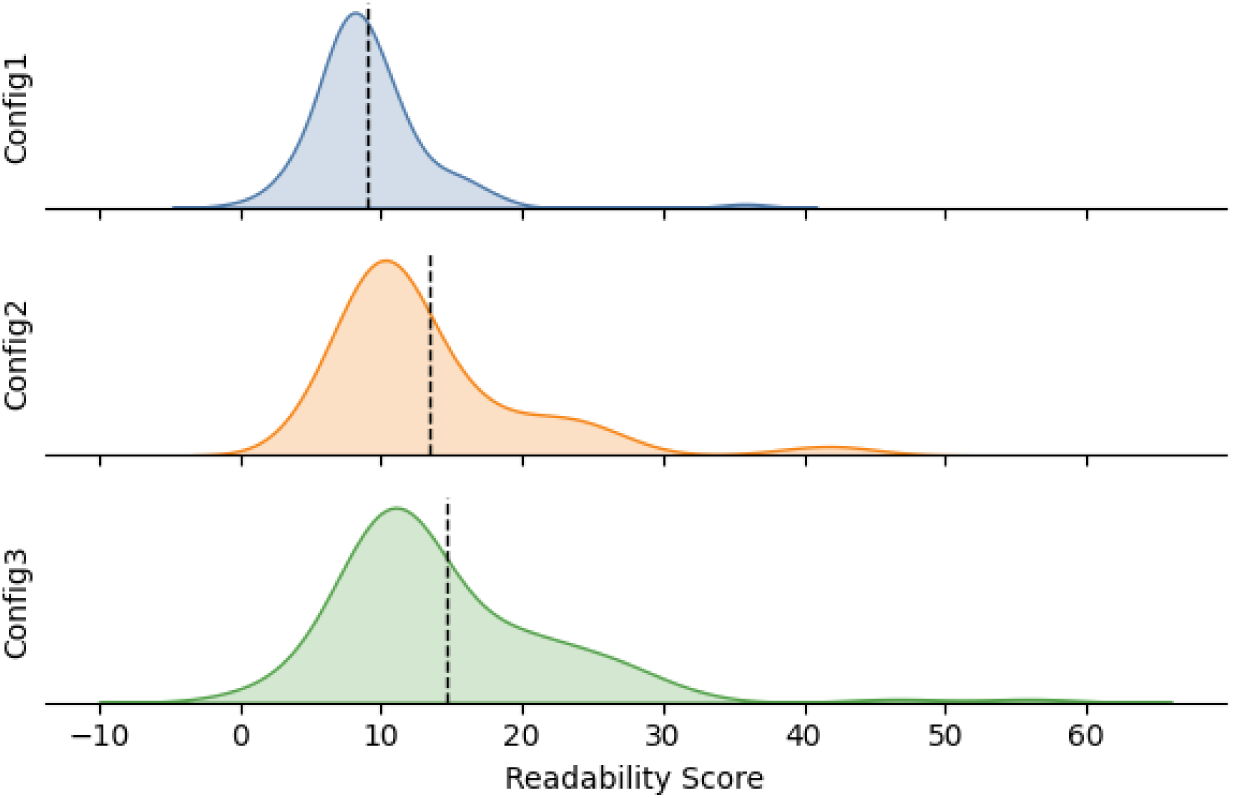
Readability score distribution over the 3 configurations.

**Figure 4.**
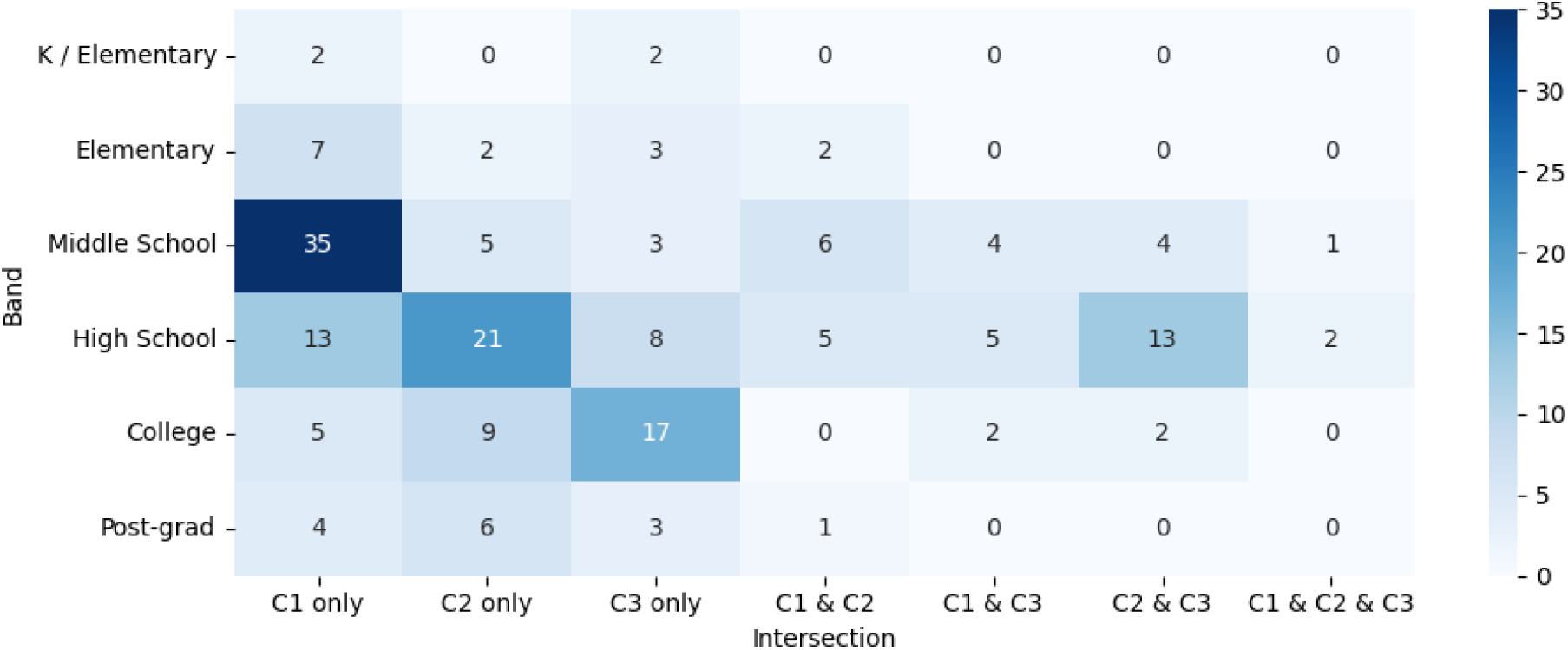
Readability score distribution over the readability score bands.

### Accuracy

Information accuracy scores for the evaluated LLMs on DiaBench are detailed in Figure 5. As mentioned above, each question was prompted three times, and the majority vote from the tested LLM was used as the final solution. On configuration 1, most models, including Gemini and GPT-5, achieved high accuracy (≥98.5%). However, the Perplexity (90.6%) and Mistral (66.7%) models posted lower scores. In configuration 2, Mistral’s performance remained significantly lower than its peers’. Interestingly, the Perplexity model, which underperformed on the seemingly easier set of configuration 1 (Config 1), achieved top-tier accuracy (99%) on the configuration 2 questions, comparable to other leading models. For configuration 3, nearly all models exhibited some performance degradation. Across all evaluations, Mistral performed significantly worse than other models, while OpenAI GPT-5 and DeepSeek-R1 are ranked among the top performers.

**Figure 5.**
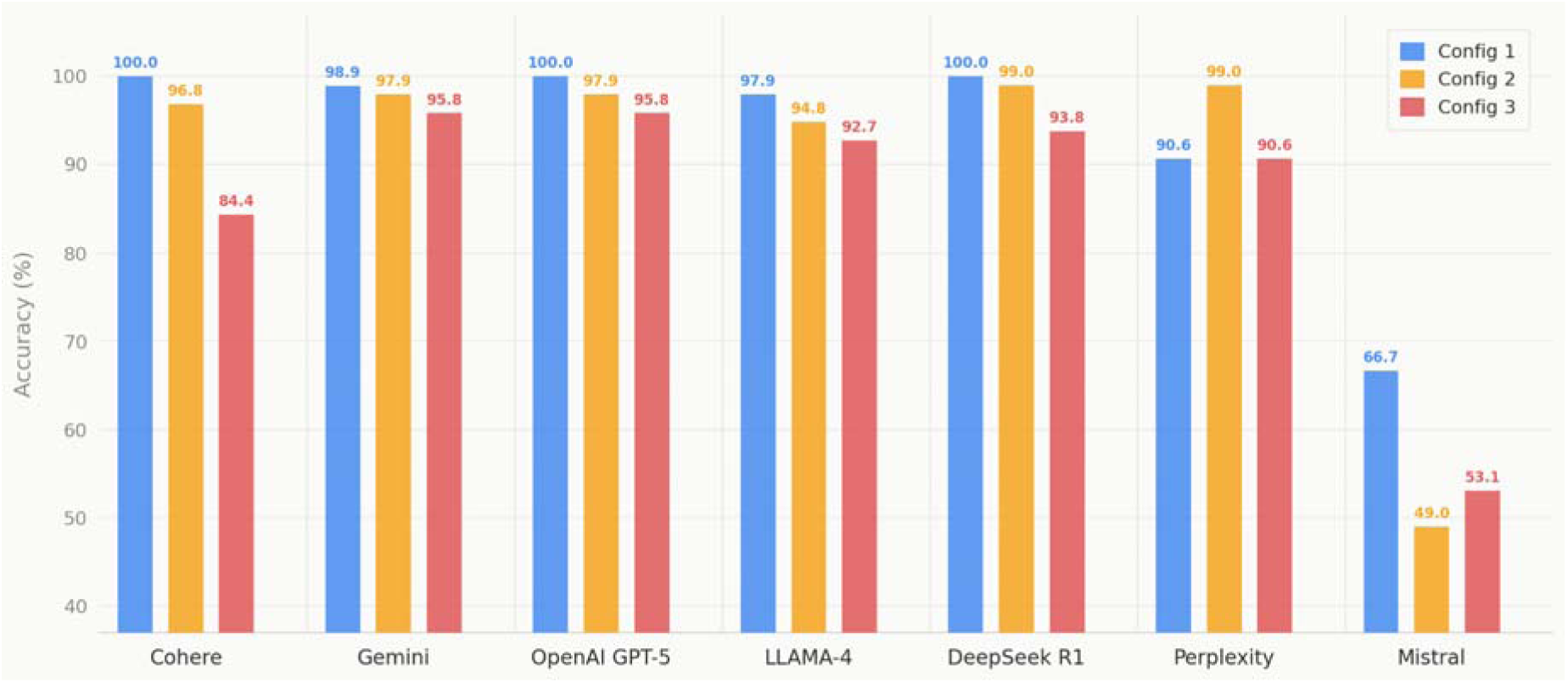
Comparison of accuracy across selected LLMs on the DiaBench MCQs across three configurations.

### Self-consistency

In Figure 6, we further examined LLM self-consistency on DiaBench. We measure self-consistency by whether we obtain the same output across all three independent queries. As anticipated, the self-consistency score decreased with increasing configuration complexity. Cohere, Gemini, and GPT-5 maintained high self-consistency on DiaBench MCQs, whereas Perplexity and Mistral demonstrated a higher propensity for nondeterministic behavior and received the lowest self-consistency scores.

**Figure 6.**
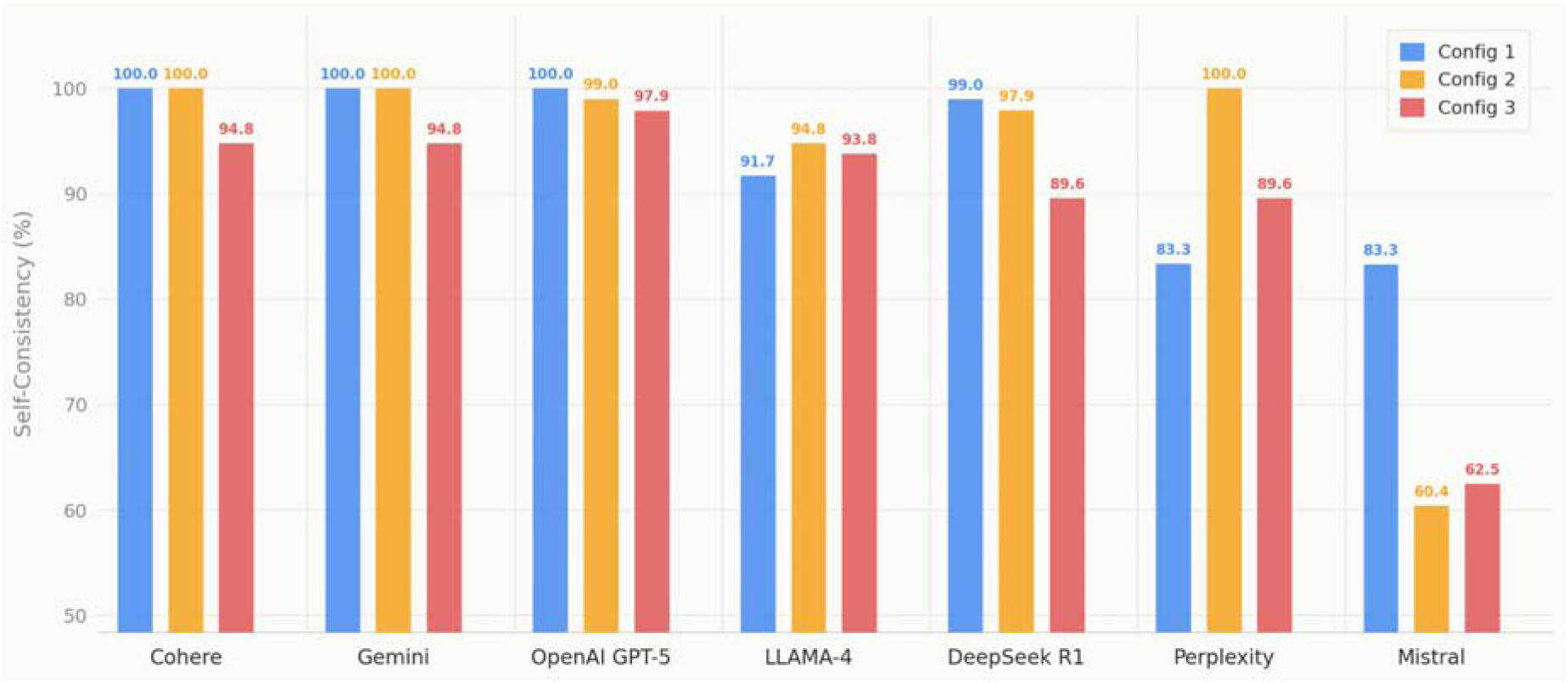
Comparison of self-consistency across selected large language models (LLMs) on the DiaBench MCQs across three configurations.

## Discussion

### Expert Verification Choices and Error Analysis

Qualitative analysis of the expert verifications revealed specific patterns in model transformation errors. As the results above show, not all questions are perfectly transformed. Issues involving inappropriate question rephrasing were more prevalent in configuration 2. First, experts observed that the model frequently introduced unprompted clinical context into the generated questions from the original Q&A.

For example, an original question,

> *“What liver test is recommended for people with diabetes or pre-diabetes?”*

was transformed into

> *“A person with diabetes has a FIB-4 score of 1.3 or higher. Which test is recommended to check for liver scarring (fibrosis)?”*

It introduces the FIB-4 score, which was not present in the source material.

Furthermore, reviewers noted that the rephrased items adopted a more formal, “test-like” tone, a characteristic particularly evident in configuration 2. Regarding the generated answer options, flagged issues primarily involved overlapping or poorly differentiated distractors (e.g., generating options such as “Over $400 billion in 2022” alongside “About $800 billion in 2022”). Some questions, like “*Which option best reflects the typical medication approach for managing type 2 diabetes, including the usual first-line drug and common add-on choices based on individual needs?*” were flagged because the generated clinical scenarios or distractors deviated slightly from standard daily clinical workflows or current best-practice guidelines. Note that the verifications are primarily done on configurations 1 and 2, as those are most likely to appear in real-world clinical encounters. Correctness of Q&As in Configuration 3 was confirmed for evaluation. This literacy configuration transformed the original Q&As dramatically by introducing lengthy and nuanced context, which is more likely to happen in personal counseling encounters rather than a generic patient educational scenario. Nevertheless, this configuration is still valuable for benchmarking purposes as it has shown greater discriminative power of ranking models and lower saturation for guiding future model improvements towards precision diabetes education.

### Generalizability to Other Clinical Settings and Specialties

There are a few things we would like to highlight for DiaBench. First, the MCQ transformation pipeline, evaluation pipeline, and the AI & clinician co-designed workflow are open-source, highly reusable, and easily generalizable beyond T2D education. This pipeline only requires high-quality Q&A data as input to conduct a comprehensive LLM evaluation of selected clinical specialties and thus empower medical practitioners to perform clinically relevant and quantitative LLM evaluation, instead of relying on generic benchmark results for model selection. While an expert verification process could further enhance the power of such evaluations, our results show that strong LLMs can generally perform the MCQ transformation with high fidelity and greatly reduce the expert verification burden. Overall, we encourage future work to reuse and extend our pipeline to many other clinical settings and specialties.

### Recommendations Over Choice of LLMs

Our aggregate analysis of both accuracy and self-consistency identifies Gemini, GPT-5, and DeepSeek-R1 as the top-performing models. These models consistently demonstrated superior accuracy and robust self-consistency, required for patient education applications. While most models perform reasonably well on Config 1’s questions, our findings also highlight a critical limitation: even state-of-the-art models struggle with questions that demand multi-step reasoning or complex analogies. Enhancing the deep clinical reasoning capabilities and generative self-consistency of LLMs remains an urgent avenue for future research.

### Limitations and Future Work

**Q&A Coverage:** While the initial Q&A set covers a wide range of daily interactions between clinicians and patients and aligns with diabetes care guidelines, it may require regular updates to reflect annual updates to ADA guidelines ^48–50^. Future iterations of DiaBench should also augment the benchmark with a larger volume of Q&A datasets, preferably with more personalized context.

**Zero-Shot Prompting Template:** All of our experiments use zero-shot prompting^51^ (i.e., without any examples). Recent research indicates that advanced prompting strategies, such as chain-of-thought reasoning and agentic frameworks, can enhance instruction-following and task performance^51–53^. Future work should explore how varying prompt strategies and agentic architectures may impact LLM accuracy and self-consistency within the DiaBench framework.

## Conclusion

In this study, we developed and validated DiaBench, an AI & clinician co-designed reusable workflow and benchmark for quantitatively assessing LLM performance in T2D diabetes education. Building on 96 representative Q&As collected from real-world encounters by CDCESs, MDs, and endocrinologists, we evaluated 7 state-of-the-art LLMs across three configurations. While models such as Gemini, GPT-5, and DeepSeek-R1 demonstrated strong accuracy and self-consistency in configurations 1 and 2, the performance degradation observed in configuration 3 highlights the necessity for domain-specific evaluation prior to the clinical deployment of artificial intelligence in patient-facing educational roles.

## Data Availability

We will open-source our code and data. Initial collected questions, generated MCQs, and code are available at https://github.com/theBigTao/DiaBench

## Institutional Review Board Statement

Institutional Review Board approval was not sought because this study did not involve human subjects research as defined under applicable institutional and regulatory guidelines.

## Author Contributions

YL: Data curation, Formal analysis, Investigation, Methodology, Software, Visualization, Writing – original draft, Writing – review and editing. Syna: Data curation, Formal analysis, Investigation, Methodology, Software, Visualization, Writing – review and editing. CYZ: Data curation, Formal analysis, Investigation, Methodology, Software, Visualization, Writing – review and editing. BK: Data curation, Investigation, Methodology, Visualization, Writing – review and editing. EH, AFS, MMS, HM: Investigation, Writing – review and editing. AS, YKL, JK, MB, DCK: Data curation and Validation, Investigation, Methodology, Writing – review and editing. MPS: Conceptualization, Supervision, Writing – review and editing. HPZ: Study design, Methodology, Supervision, Writing – review and editing. TW: Conceptualization and study design, Data curation, Methodology, Project administration, Supervision, Writing – review and editing.

## Acknowledgements

EH is supported by T32HD040128 from the NICHD/NIH. The content is solely the responsibility of the authors and does not necessarily represent the official views of the National Institutes of Health.

## Appendix

## Appendix A: Flesch-Kincaid Grade Level Details

Flesch-Kincaid grade level is split into 6 levels: kindergarten / elementary (score: 0-3), elementary (score: 3-6), middle school (score: 6-9), high school (score: 9-12), college (score: 12-15), and post-grad (score:15-18). More details can be found in the following link. https://readable.com/readability/flesch-reading-ease-flesch-kincaid-grade-level/

## Appendix B: MCQs Transformation Prompt for 3 Configurations

The exact prompt templates used in the MCQ transformation from Q&A are attached below. The major differences between configurations are **bolded**.

### Configuration 1

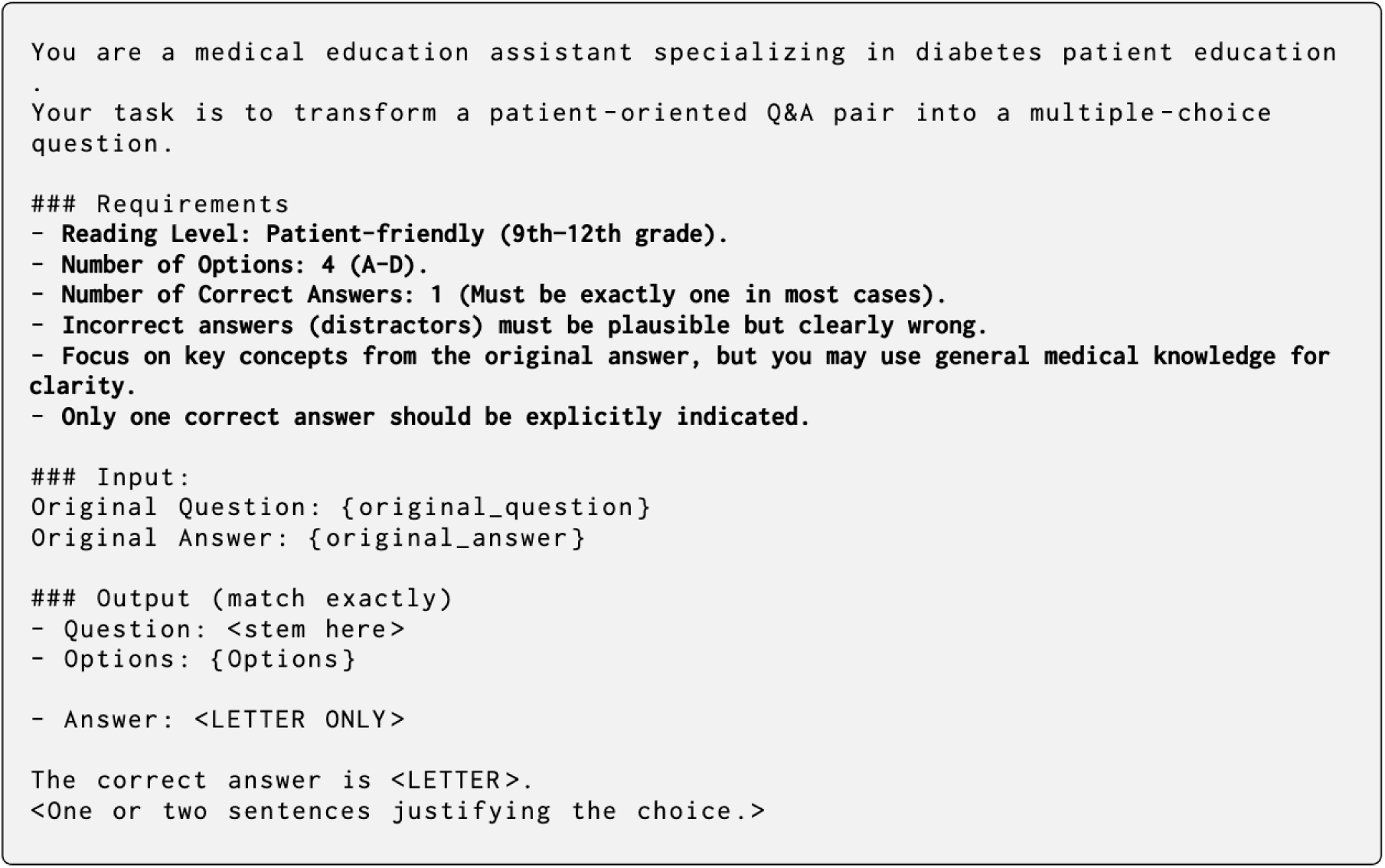

### Configuration 2

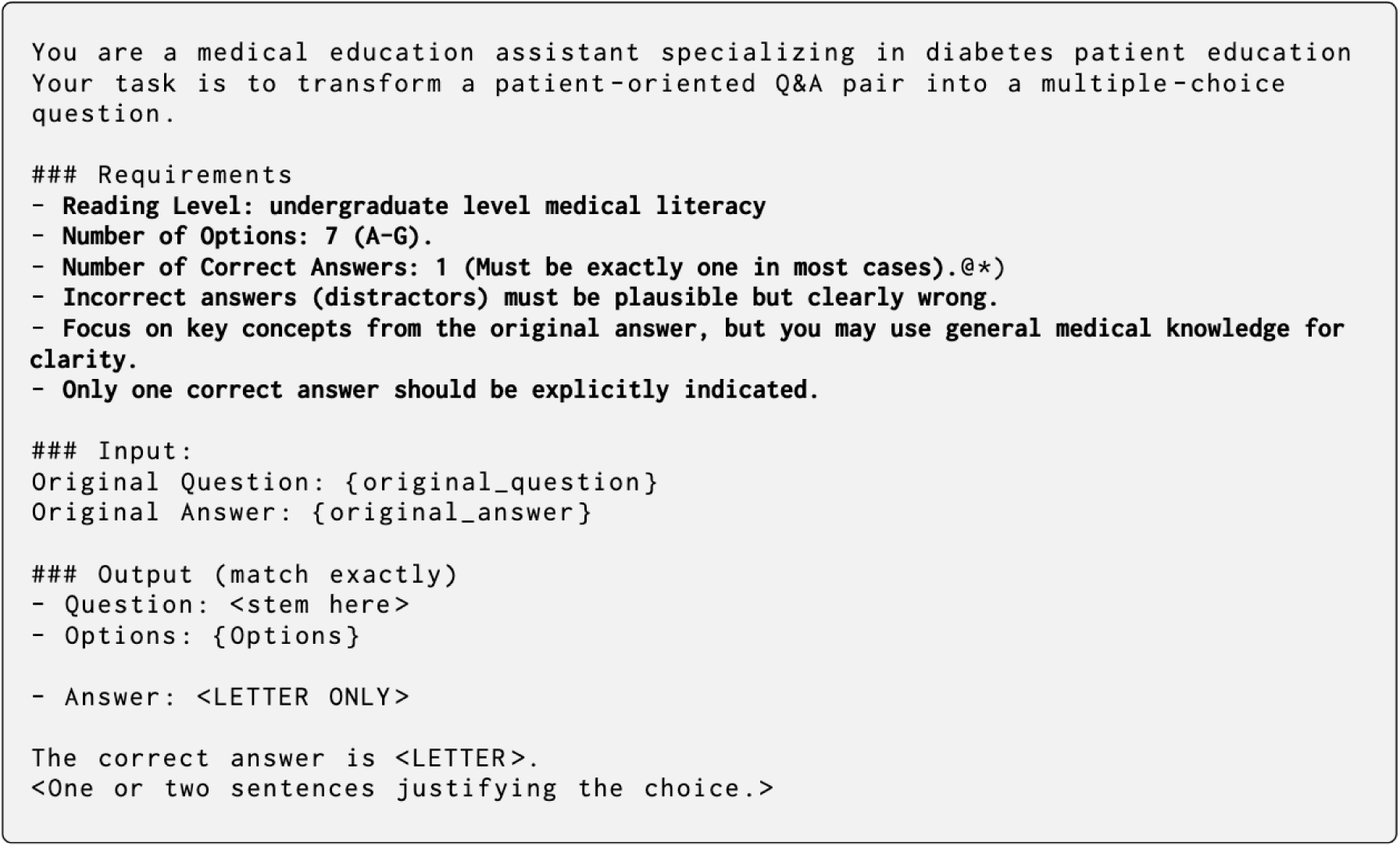

### Configuration 3

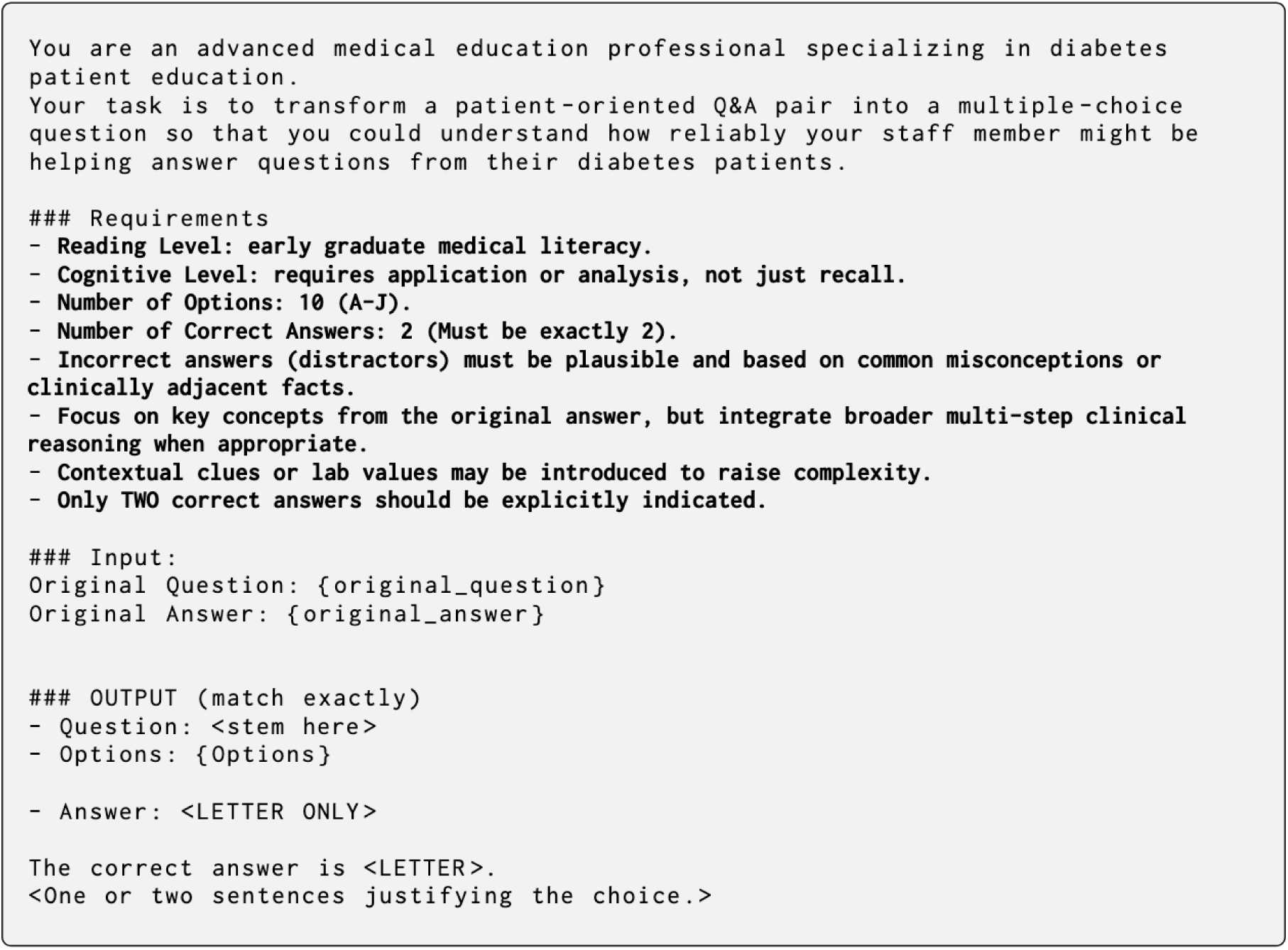

